# [^11^C]Carfentanil PET Whole-Body Imaging of Mu-Opioid Receptors: A First In-Human Study

**DOI:** 10.1101/2024.12.31.24319819

**Authors:** Jacob G. Dubroff, Chia-Ju Hsieh, Corinde E. Wiers, Hsiaoju Lee, Elizabeth J. Li, Erin K. Schubert, Robert H. Mach, Henry R. Kranzler

**Affiliations:** Department of Radiology, Perelman School of Medicine, University of Pennsylvania, Philadelphia, PA 19104, United States; Department of Psychiatry, Perelman School of Medicine, University of Pennsylvania, Philadelphia, PA 19104, United States; Mental Illness Research, Education and Clinical Center, Crescenz VAMC, Philadelphia, PA 19104, United States

**Keywords:** Mu-opioid receptors, [^11^C]carfentanil, naloxone, positron emission tomography, PennPET Explorer, reference region, sex differences, long axial field-of-view

## Abstract

**Introduction:** Mu-opioid receptors (MORs) are G-coupled protein receptors with a high affinity for both endogenous and exogenous opioids. MORs are widely expressed in the central nervous system (CNS), peripheral organs, and the immune system. They mediate pain and reward and have been implicated in the pathophysiology of opioid, cocaine, and other substance use disorders. Using the long axial field-of-view (LAFOV) PennPET Explorer instrument and the MOR selective radioligand [^11^C]carfentanil ([^11^C]CFN), we measured the ***whole-body*** distribution of MORs in 13 healthy humans. We also examined sex differences in MOR distribution at baseline and after pretreatment with the MOR antagonist naloxone.

**Methods:** Six female and seven male healthy subjects underwent two [^11^C]CFN PET imaging sessions—one at baseline and one immediately following pre-treatment with the MOR antagonist naloxone (13 mcg/kg). Whole-body PET imaging was performed on the PennPET Explorer, a 142-cm axial bore instrument. [^11^C]CFN brain distribution volume ratios (DVRs) were determined using the occipital cortex and the visual cortex within it as reference regions. For peripheral organ DVRs, the descending aorta and proximal extremity muscle (biceps/triceps) were used as reference regions.

**Results:** Naloxone blockade reduced MOR availability by 40-50% in the caudate, putamen, thalamus, amygdala, and ventral tegmentum, brain regions known to express high levels of MORs. Women showed greater receptor occupancy in the thalamus, amygdala, hippocampus and frontal and temporal lobes and a greater naloxone-induced reduction in thalamic MOR availability than men (p’s <0.05). For determining brain MOR availability, there was less variance in the visual cortex than the occipital cortex reference region. For peripheral MOR determination, the descending aorta reference region showed less variance than the extremity muscle, but both showed blocking effects of naloxone.

**Conclusions:** [^11^C]CFN whole-body PET scans are useful for understanding MOR physiology under both baseline and blocking conditions. Extra-CNS reference regions may be useful for quantifying radiotracers when a region devoid of binding in the CNS is unavailable. The LAFOV PET instrument was useful for measuring changes in the short-lived radiotracer [^11^C]CFN, with and without naloxone blocking. Further research is needed to evaluate the behavioral and clinical relevance of sex differences in naloxone-MOR interactions.

## Introduction

Opioid misuse is a worldwide epidemic associated with high overdose and mortality rates (*1,2*). The mu-opioid receptor (MOR) is the target of opioid drugs of abuse including fentanyl and heroin, as well as methadone and buprenorphine, opioid agonist medications used to treat opioid use disorder (OUD). A major risk associated with MOR agonists is respiratory depression (*3*). Naloxone is a highly effective, short-acting opioid antagonist that is widely used to reverse the effect of opioids (*4,5*), and in combination with buprenorphine for the chronic management of OUD (*6–8*). A better understanding of the interactions of agonists and antagonists with the MOR could help to elucidate the etiology, prevention, and treatment of OUD.

MORs are widely expressed in the central nervous system (CNS), peripheral organs, and immune system (*9,10*). Although MOR pharmacology has been extensively studied using positron emission tomography (PET) imaging with radioligands such as [^11^C]carfentanil ([^11^C]CFN) *in vivo* in humans (*11–15*), non-human primates (NHPs) (*16,17*), and rodents (*18*), these observations have largely been restricted to the brain in humans and NHPs due to the limits of available instrumentation.

### [^11^C]CFN-PET imaging has advanced our understanding of MOR behavior

Carfentanil, a potent MOR agonist with abuse potential, was developed for use in veterinary medicine (*19,20*). [^11^C]CFN-PET is a short-lived C-11 tracer that in 1989 was first used to measure MOR availability (*21*). It has subsequently been used to examine the role of the MOR in pain (*22–26*), and to study a variety of pathological conditions (e.g., obesity, gambling), exercise, and the experiences of pleasure and pain (*27–30*). Alterations in MOR availability and indirectly, endorphin release, have been measured following exposure to amphetamine, nicotine, alcohol, and nicotine in a variety of populations (*29,31–35*).

Using [^11^C]CFN-PET brain imaging, MOR availability has also been shown to increase with age in neocortical areas and the putamen of both sexes (*26,36*). Although women generally have higher MOR availability in both cortical and subcortical areas (*26,36*), postmenopausal women have lower MOR availability than men in the thalamus and amygdala (*36*). Using [^11^C]CFN-PET, women showed a greater association than men between beta-endorphin release and genetic risk for OUD and major depression during exposure to a stressful stimulus (*37*), underscoring the potential clinical relevance of MOR-related sex differences. Although dynamic interactions of naloxone with MORs have been studied with [^11^C]CFN-PET in men (*7*), sex differences in naloxone binding were not examined, despite their potential clinical relevance.

### Neuroreceptor measurements using long axial field-of-view (LAFOV) PET instruments

With only two notable exceptions that examined normal cardiac and lung cancer MOR availability(*38,39*), [^11^C]CFN-PET studies have been largely limited to the brain, with no studies of the spinal cord or peripheral organs. The development of long axial field-of-view (LAFOV) PET instrumentation has enabled a new focus in molecular imaging (*40–43*). This innovative scanning technology combines the advantages of greater count sensitivity with dynamic whole-body imaging (*40*). A prototype instrument, the PennPET Explorer was developed by the Physics and Instrumentation group at the University of Pennsylvania (*41–44*), which enabled the first whole-body C-11 radiotracer imaging of NHPs using [^11^C]CFN (*45*). Such instruments may allow for reference region methods for tracers that do not have a valid brain region. Furthermore, the PennPET Explorer’s high temporal resolution could be harnessed to examine arterial input functions needed to measure kinetic parameters and improve PET quantitation approaches. Here, we present the first human whole-body neuroreceptor PET imaging study using [^11^C]CFN where extra-CNS reference regions (aorta and muscle) are investigated.

## Material and methods

### Participants

This study was conducted in accordance with the Declaration of Helsinki and all procedures were approved by the University of Pennsylvania’s Institutional Review Board and the study registered on clinicaltrials.gov (NCT05528848). Healthy, English-speaking individuals aged 18-45 gave informed consent to participate in the study. Using the Mini International Neuropsychiatric Interview(*46*), we excluded prospective participants with a history of Diagnostic and Statistical Manual of Mental Disorders, 5^th^ edition (*47*) psychiatric or substance use disorders. We also excluded individuals taking psychotropic medications, those who were pregnant or planning to become pregnant, or breastfeeding; had a history of brain injury or material in the body that contraindicates magnetic resonance imaging (MRI). Subjects underwent urine drug testing at both the baseline visit and on the day of PET imaging to ensure the absence of an exogenous substance that could interfere with testing procedures.

### MRI Imaging

To facilitate automatic generation of brain volumes of interest (VOIs), all participants received a T1-weighted anatomic MRI scan on a 3-T Prisma Fit Siemens scanner prior to the first of the two PET scans.

### Preparation of [^11^C]CFN

[^11^C]CO_2_ was produced using the IBA cyclotron at the University of Pennsylvania by a ^14^N (p,α) ^11^C reaction on an 0.5% O_2_ in N_2_ gas target. Briefly, [^11^C]CO_2_ was trapped in a liquid nitrogen-cooled tube in the Synthra MeIPlus synthesis module (Synthra GmbH, Germany). [^11^C]CO_2_ was converted to [^11^C]CH_4_ using a molecular sieve, nickel and hydrogen. [^11^C]CH_4_ was converted to [^11^C]CH_3_I using iodine in a heated loop and bubbled into the reaction mixture containing desmethylcarfentanil, dissolved in DMF. The solution was heated to 60°C for 5 min. The resulting radiolabeled products were purified with a C2 Bond Elut cartridge (Agilent, US) after dilution with 1% ammonium hydroxide solution. The product was eluted with ethanol and washed with sterile saline. The final product formulation was filtered through a 0.22 μm filter before being collected in a sterile final product vial. The finished products were tested for chemical and radiochemical purity by HPLC analysis. The specific activity was 480.5±401.2 MBq/nmol.

### [^11^C]CFN PET data acquisition

All [^11^C]CFN PET scans were conducted on the PennPET Explorer(*42,43*). A low-dose CT scan was performed for anatomical localization and attenuation correction followed by a 90- min dynamic PET image acquired in list-mode after venous injection of 168.5 ± 58.6 MBq (injected mass: 19.6±8.4 ng/kg) of [^11^C]CFN. All PET images were reconstructed using a time-of-flight list-mode ordered subsets expectation maximization (OSEM, 25 subsets) reconstruction algorithm(*43*). The reconstructed images had a matrix size of 300×300×712 and a voxel size of 2×2×2 mm.^3^

### Image analysis

All whole-body PET/CT images were processed and analyzed using PMOD software (version 4.2, PMOD Technologies Ltd., Zurich, Switzerland). Each perfusion phase (1-10 min) was cropped to focus on the brain and then was co-registered to the corresponding T1 image.

Individual T1 images were spatially normalized to the Montreal Neurologic Institute (MNI) T1 brain template(*48*). The spatial normalization parameters were applied to the corresponding dynamic brain PET data to transfer the images to MNI space. Five bilateral volumes of interest (VOIs) were determined using the Automated Anatomical Labeling (AAL) atlas (*49*) for analysis: bilateral amygdala, caudate, putamen, thalamus, and ventral tegmentum. The VOI was determined manually for the ventral tegmentum. The distribution volume ratio (DVR) for each brain VOI was computed with the Logan reference tissue model (*50*) using the visual cortex (modified from calcarine) as the reference tissue with fixed k2’ (0.1237 min^-1^) from the literature (*12,21*). The DVR was calculated using a 0-90 min time-activity curve (TAC). The percent receptor occupancy (%RO) of the blocking effect for the naloxone pretreatment study was calculated using the following equation:

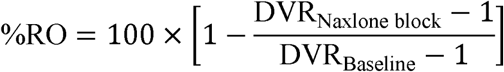

Additional VOIs of spinal cord (cervical, thoracic, and lumbar), spinal bone marrow, and peripheral organs (aorta, heart wall, kidneys, spleen, small intestine, spinal bone marrow (C-, T-, and L-), muscle, liver, and stomach were manually delineated on the PET or CT image of each scan. Area under the curve (AUC), summed standard uptake value (SUV), and standard uptake value ratio (SUVR_Aorta_) analysis using data acquired 50-70 min following [^11^C]CFN administration were also used to measure radiotracer distribution. Prior work by Madar et al. examining [^11^C]CFN in primary non-small cell lung cancer supports these approaches (*38*).

### Statistical analysis

Statistical analyses were performed using SPSS version 29.

### Brain MOR availability

<u>DVRs:</u> Brain DVRs were analyzed using repeated-measures ANOVAs (5 VOIs: thalamus, caudate, putamen, amygdala, and ventral tegmentum using visual cortex as the reference region), with the naloxone intervention (baseline/naloxone) as a within-subject factor and sex (female/male) as a between-subject factor. A repeated-measures ANOVA was also performed on the %RO, with sex as a between-subject factor. *Post hoc* two-tailed P-values < 0.05 were considered statistically significant.

### Whole-body MOR availability and reference regions

<u>SUVs and SUVRs:</u> Paired t-tests were performed on whole-body SUVs and SUVR_Aorta_ (50-70 min post injection interval) for the following 14 peripheral regions: aorta, heart wall, kidneys, spleen, small intestine, spinal bone marrow (C-, T-, and L-), spinal cord (C-, T-, and L-), muscle, liver, and stomach. Bonferroni-corrected p-values of p<0.0036 (0.05/14) were considered statistically significant.

Paired t-tests were also used to compare the baseline and the naloxone conditions on the AUC of the TACs for the occipital cortex, as defined by the AAL atlas and the visual cortex, a region defined as calcarine cortex in the AAL atlas. Finally, paired t-tests were performed between baseline and naloxone on the AUC of the TACs for aorta, muscle, cervical spinal cord, thoracic spinal cord and lumbar spinal cord.

## Results

### Brain MOR availability

Participant characteristics and the [^11^C]CFN dose administered during baseline and naloxone scans are shown in Table 1.

**Table 1:** Participant Characteristics.

| Scans | All | Male | Female |
| --- | --- | --- | --- |
| Sample Size | 13 | 7 | 6 |
| Scans | 26 | 14 | 12 |
| Age | $29.1 \pm 8.3$ | $29.4 \pm 7.4$ | $28.7 \pm 10.1$ |
| Weight | $74.5 \pm 16.7$ | $82.8 \pm 19.3$ | $64.9 \pm 3.6$ |
| Injection Dose (MBq) |  |  |  |
| Baseline | $167.1 \pm 79.6$ | $178.0 \pm 81.0$ | $154.3 \pm 83.4$ |
| Naloxone Blocking | $164.3 \pm 32.3$ | $170.7 \pm 32.7$ | $156.7 \pm 33.0$ |

<u>DVR</u>: Representative MOR availability at baseline and following naloxone blockade using the visual cortex as a reference region is shown in Figure 1. Visual cortex baseline and naloxone pretreatment TACs were less distinguishable when compared to the occipital lobe (Supplemental Figure S1) due to the automated definition of that region, which includes spillover counts in adjacent structures. There was a significant main effect of the naloxone intervention on the [^11^C]CFN Logan DVRs (F_1,11_=1132.9, p<0.001), with lower DVRs following naloxone than at baseline for all brain VOIs (all p<0.0001). The effects of VOI (F_4,8_=31.7, p<0.001) and the interaction of naloxone x VOI (F_4,8_=35.1, p<0.001) were also significant (Figure 2).

**Figure 1.**
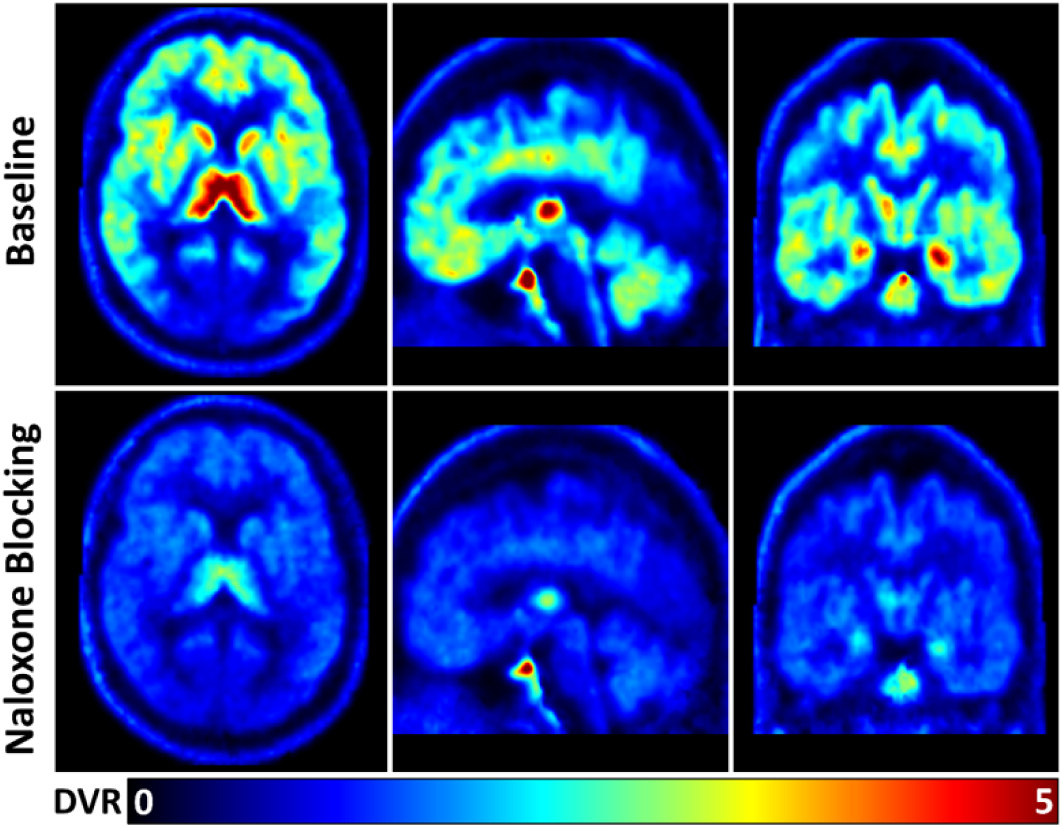
DVR-weighted [^11^C]CFN-PET brain images for a 42-year-old, healthy male at baseline (top row) and following pretreatment with 13 mcg/kg naloxone (bottom row) using the visual cortex as a reference region. Images are shown in the transaxial (left column), sagittal (middle column), and coronal (right column) planes. A DVR intensity scale is at the bottom of the panel.

**Figure 2.**
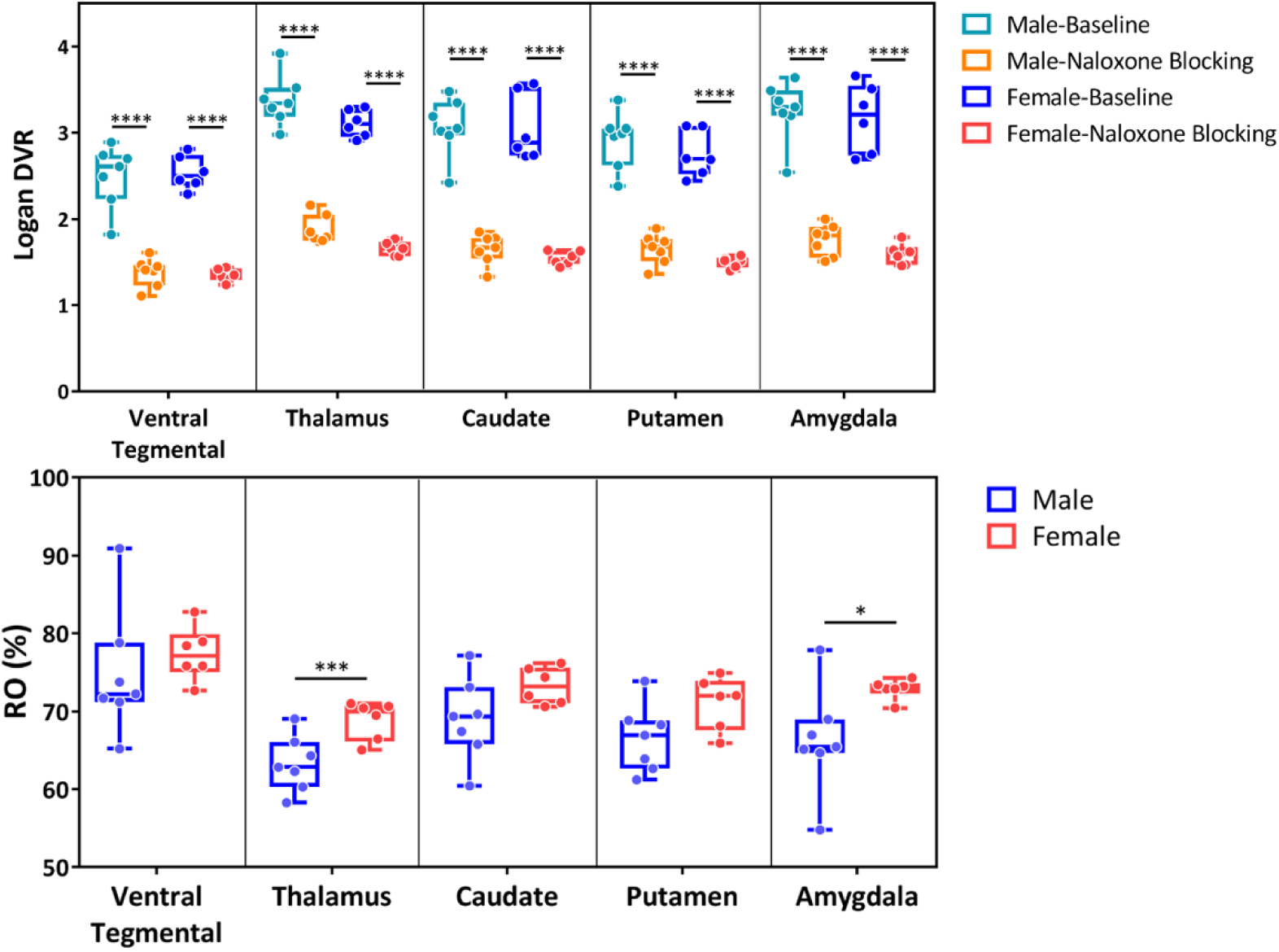
Brain [^11^C]CFN DVR in MOR rich regions reveal robust blockade of MOR availability with naloxone (top) and sex differences in % MOR receptor occupancy for the thalamus and amygdala (bottom). * p<0.05, *** p<0.001, ****p<0.0001.

There were no effects of sex on baseline MOR (F_1,4_=1.6, p=.23), intervention x sex (F_1,11_=0.2, p=0.65), or intervention x VOI x sex (F_4,8_=2.1, p=0.17) (Figure 2). Exploratory analyses showed that thalamic DVRs were non-significantly lower in women than men both at baseline (p=0.074) and after naloxone (p=0.014).

<u>Naloxone %RO</u>: There was a significant main effect of sex on naloxone %RO (F_1,11_=5.2, p=0.043), with %RO in the thalamus (p=0.009) and amygdala (p=0.040) greater among women than men. There were also non-significant sex difference in the caudate (p=0.094), putamen (p=0.062), and VTA (p=0.49). There was a significant main effect of VOI on %RO (F_4,8_=10.6, p=0.003), but no interaction of VOI x sex (p=0.94). There was no effect of age on %RO (F_1,11_=0.12, p=0.73). Sex differences for DVRs in MOR-rich regions at baseline and after naloxone, and in naloxone %RO are shown in Figure 2. Supplemental Figure S3 shows the %RO for the naloxone condition in all analyzed brain regions. Supplemental Table T1 includes all tested DVRs and %RO data by sex.

### Comparison of central versus peripheral reference regions

TACs for the three reference regions (visual cortex, descending aorta, and upper extremity muscle) are shown in Figure 3. The AUC was significantly reduced with naloxone pre-treatment for the visual cortex (t_12_=3.92, p=0.002), but neither the aorta (t_12_=0.86, p=0.41) nor muscle (t_12_=0.46, p=0.91).

**Figure 3.**
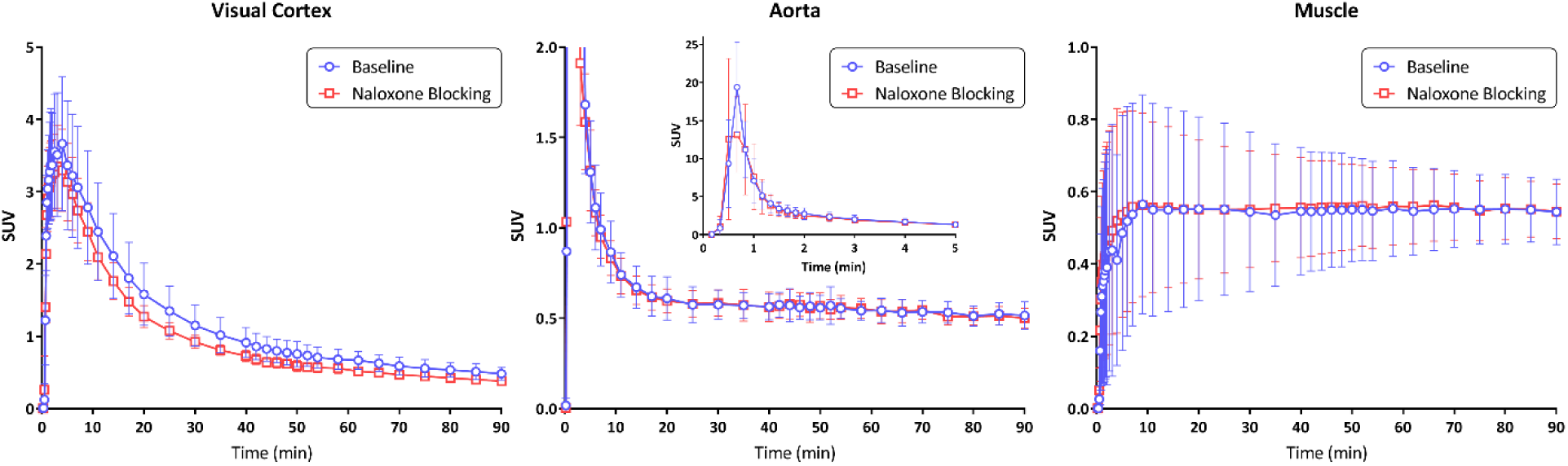
Reference TACs are shown in SUV of three reference regions: visual cortex (left panel), descending aorta (middle panel), and upper extremity muscle (right panel). Baseline curves are shown in blue and naloxone pre-treatment curves in red. Optimally, the reference region should not differ between baseline and blocking conditions.

Mean DVR values for the 5 MOR-rich regions using the established visual cortex (DVR_visual cortex_) and either the aorta (DVR_aorta_) or muscle (DVR_muscle_) as comparators showed linear relationships (Figure 4) albeit with a positive bias (slope=1.54 and 2.06, R=0.8373 and 0.6889, respectively). Use of the descending aorta (SUVR_aorta_) or upper extremity muscle (SUVR_muscle_, triceps/biceps) yielded robust measures of MOR availability following naloxone blockade in MOR-rich brain regions (see Supplemental Figure S2). Irrespective of the reference region (visual cortex, aorta, or muscle), differences in DVR between baseline and naloxone blockade were statistically significant (p<0.0001) in the 5 MOR-rich regions: ventral tegmentum, thalamus, caudate, putamen, and amygdala. SUV, SUVR_aorta_, or SUVR_muscle_ approaches all normalize [^11^C]CFN-PET data in similar distributions. Representative data are shown in Figure 5.

**Figure 4.**
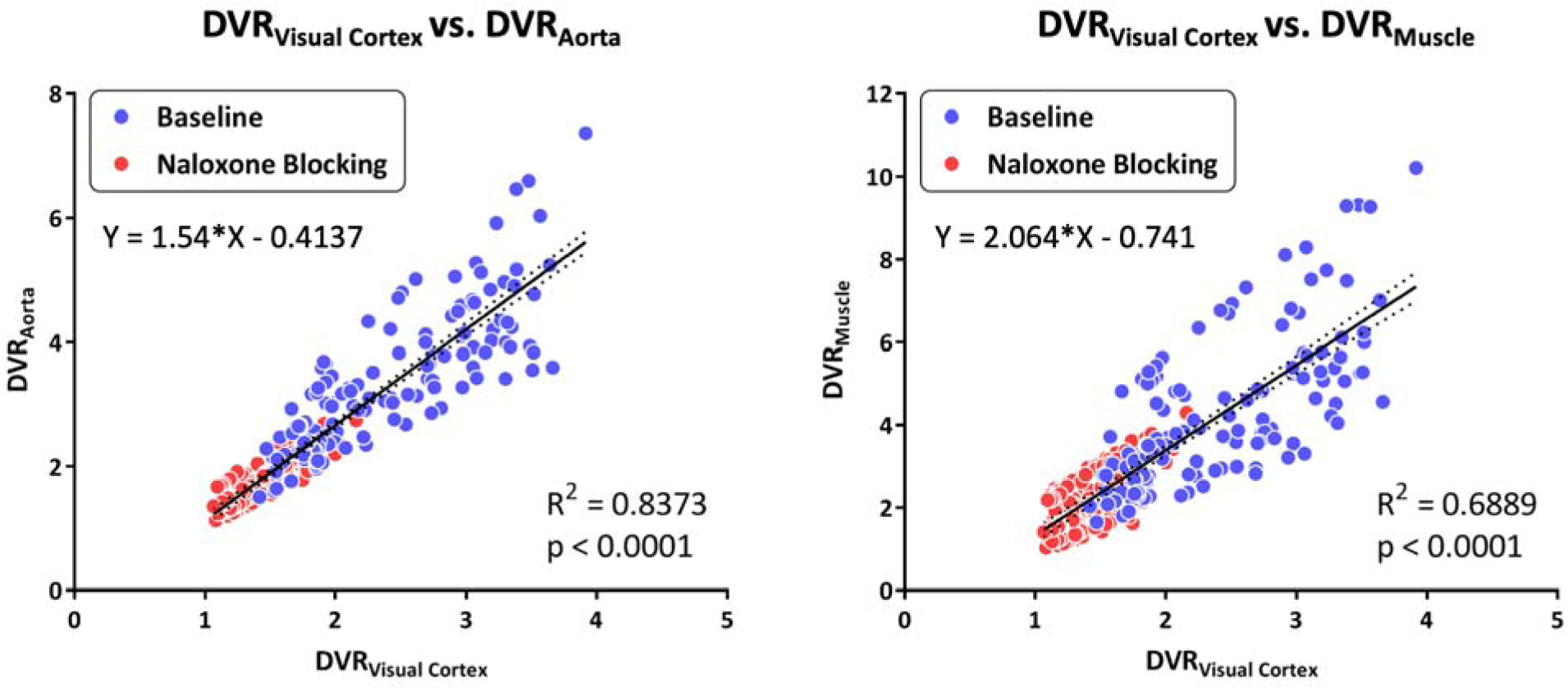
There is a linear relationship between the established DVR_visual cortex_ with either DVR_aorta_ or DVR_muscle_. The left panel of figure 3 shows DVR_Aorta_ plotted DVR_visual cortex_ at both baseline (blue) and naloxone blocking (red) conditions. The right panel shows DVR_aorta_ plotted DVR_muscle._ DVR_aorta_ more strongly correlates to DVR_visual cortex_ than DVR_muscle_ (R^2^=0.8373 and 0.6889, respectively). Dotted lines represent 95% confidence range for the solid linear regression line.

**Figure 5.**
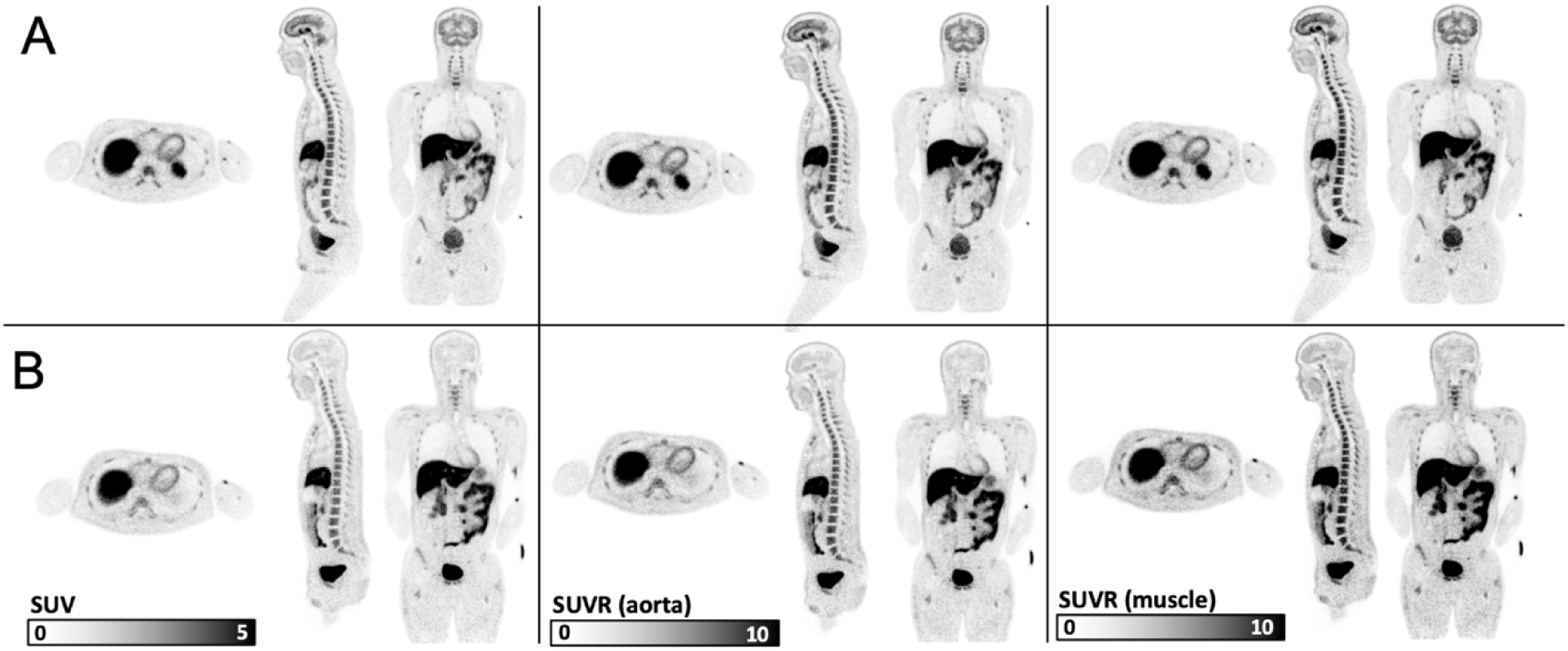
Strategies for whole body semi-quantification of [^11^C]CFN-PET distribution. This figure Summed images acquired 50-70 min post-injection of a representative subject at baseline (A, top row) and following naloxone administration (B, bottom row). The three columns represent different quantification strategies: SUV sum (first column), SUVR with aorta as the reference region (second column), and SUVR with upper extremity muscle as the reference region (third column). Note that the naloxone blocking effect in brain is evident using each approach.

Exploratory analyses also revealed a significant difference in AUC with naloxone pre-treatment for the occipital cortex (t_12_=5.43, p<0.001), the cervical spinal cord (t_12_=8.0, p<0.0001), and thoracic spinal Cord (t_12_=3.6, p=0.004), but not the lumbar spinal cord (t_12_=1.14, p=0.19) (Supplemental Figures S1 and S4, Supplemental Table T3).

### Whole-body [^11^C]CFN distribution

Summed SUVR DVR (SUVR_Aorta_ Figure 6), SUV, and TAC AUC (Supplemental Figure S4) approaches all showed robust differences in the PET [^11^C]CFN signal between baseline and blocking conditions in the cervical and thoracic portions of the spinal cord. Note that data are included in Supplemental Tables T2 (SUV whole-body analysis) and T3 (TACs for the cervical, thoracic, and lumbar portions of the spine). Graphs of TACs are shown in supplemental Figure S4.

**Figure 6.**
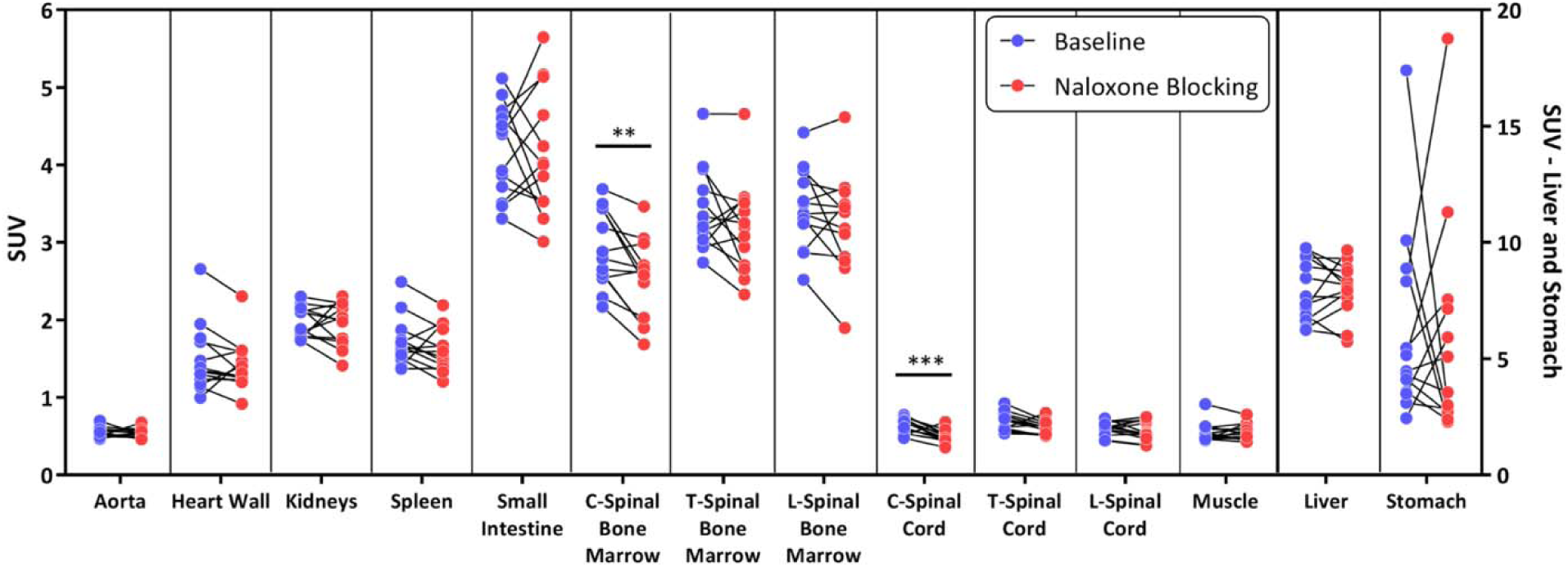
Whole body naloxone alters [^11^C]CFN distribution. This figure illustrates the effect of naloxone blockade throughout the body using the LAFOV instrument. SUVR_Aorta_ summation between 50-70 minutes following [^11^C]CFN administration are shown (* p<0.05, ** p<0.01, *** p<0.001).

Paired sample t-tests showed significant reductions in baseline SUV following naloxone in the cervical spinal bone marrow (t_12_=3.9, p=0.002), cervical spinal cord (t_12_=4.8, p=0.0004), and thoracic spinal cord (t_12_=2.6, p=0.025). The effects in the cervical spinal bone marrow and cervical spinal cord survived Bonferroni correction for 14 comparisons (i.e., both p’s<0.0036). Furthermore, paired sample t-tests also demonstrated reductions in the SUVR_Aorta_ with naloxone vs. baseline for the cervical spinal bone marrow (t_12_=3.1, p=0.010), cervical spinal cord (t_12_=3.8, p=0.003), and thoracic spinal cord (t_12_=3.0, p=0.011). Only the effect in the cervical spinal cord survived Bonferroni correction for 14 comparisons.

## Discussion

This report describes the first whole-body neuroreceptor PET imaging study in humans. [^11^C]CFN is an established radiotracer that has been used extensively at multiple institutions (*7,21,45,51*). We used the MOR agonist [^11^C]CFN and the MOR antagonist naloxone to visualize the distribution of the MOR receptor throughout the human body. In addition to the significant technical advances in PET instrumentation and quantification strategies reflected in our findings, we show sex differences in MOR receptor occupancy by naloxone—the MOR antagonist critical to treating opioid overdose and used together with buprenorphine to prevent intravenous administration of the MOR partial agonist when used in the maintenance treatment of OUD.

Zubieta et al. have previously shown greater brain MOR availability in healthy females than males, though in post-menopausal females, in the thalamus and amygdala it was less than that of males (*36*). Also using [^11^C]CFN-PET brain imaging, Smith et al. demonstrated greater MOR availability and activation of endogenous opioid neurotransmission in females during a high estrogen state induced via exogenous dosing of the hormone (*26*). Using [^11^C]CFN-PET in a large sample, we previously found greater associations of MOR availability with genetic risk of OUD and major depression among females than males (*37*). Here, we did not take into consideration the reproductive cycle status or estrogen state of the 6 healthy premenopausal females at the time of scanning. Nonetheless, the greater receptor occupancy in the brains of healthy females in our sample following weight-based naloxone administration is consistent with prior literature demonstrating sex differences in MOR binding with [^11^C]CFN-PET.

Use of the LAFOV (long axial field-of-view) PennPET Explorer PET instrument also permitted measurement of MOR availability and [^11^C]CFN distribution throughout the body. SUVr, SUV, and AUC methods all detected reductions in MOR availability in the cervical and thoracic portions of the spinal cord, known regions of MOR expression (*52*). Further development of novel quantification strategies to define the strengths and weaknesses of each approach are needed.

In keeping with a prior LAFOV scanner study that quantified the short-lived radiotracer [C-11]-butanol (*53*), our findings demonstrate robust reproducibility of a blocking effect using an established pharmacological challenge. Whereas LAFOV PET instruments provide many advantages over conventional PET/CT scanners, including superior sensitivity (*40,41,43*), they represent an important step forward for PET instrumentation that, as we demonstrate here, can be used for neuroreceptor studies.

The measurement of whole-body dynamic [^11^C]CFN at baseline and after naloxone pretreatment also supported the use of non-brain reference regions. Reference region strategies for quantifying PET neuroreceptor binding proposed over 30 years ago (*54*) depend on a comparable background region that assumes the same K_1_/k_2_ ratio as the target region (*55,56*).

Because there is no MOR expression in the visual cortex, it is well suited to serve as a reference region to estimate regional brain MOR availability for [^11^C]CFN-PET measurements. This strategy has been useful, for example, in quantifying dopaminergic-receptor-targeting radiotracers that are generally confined to the striatum (*57*). However, such a reference region is not available for many neuroreceptor tracers. Although ideally the background would be located within the brain, using a peripheral background region strategy could support quantification approaches for other neuroreceptor tracers without a reference region completely devoid of brain receptor expression.

Developing such a peripheral reference approach will require validation with established methods. Specifically, the high temporal resolution of the PennPET Explorer instrument offers the opportunity to determine individual study arterial input functions (AIFs)(*42,43*), often required for kinetic analysis, derived from image data only and potentially obviating arterial sampling for many radiotracers. Whole-body PET imaging that capitalizes on image-based AIFs with venous sampling could obviate invasive arterial line blood sampling while providing accurate, reproducible measurements (*58*). However, further studies that use arterial-line sampling to precisely measure kinetic parameters (K_1_,k_2_) and compare approaches are needed to validate this approach.

We conclude that [^11^C]CFN PET imaging using a LAFOV instrument has many advantages, which include visualizing changes induced by a pharmacological challenge throughout the body. LAFOV instruments may offer novel quantification strategies for measuring neuroreceptor tracers both within and outside the brain. Finally, sex differences in naloxone-MOR interactions should be further investigated to provide additional insights into opioid neuropharmacology, especially given the present opioid epidemic.

## Acknowledgments

We thank the University of Pennsylvania Physics and Instrumentation Group for the assistance they provided in conducting the PennPET Explorer image studies.

## Statements and Declarations

### Author Contributions

The study was designed by JGD and HRK. Material preparation, data collection, and analyses were performed by CJH, CW, HRK and JGD. The first draft of the manuscript was written by JGD and HRK. All authors read and approved the final manuscript.

### Data Availability

The article contains complete data used to support the findings of this study.

### Ethics Approval

University of Pennsylvania Institutional Review Board

### Consent to Participate

All subjects provided informed consent prior to study procedures.

### Consent to Publish

Not applicable.

### Assignment of Copyright

The authors hereby declare that this manuscript is original work, has not been published previously, and is not under consideration for publication elsewhere. We confirm that all necessary permission have been obtained for any copyrighted material included in the manuscript, and that the manuscript complies with all applicable copyright laws.

## References

1 Spencer MR, Miniño AM, Warner M. Drug overdose deaths in the United States, 2001–2021. *NCHS data brief.* 2022;457:1-8.

2. Spencer MR, Warner M, Cisewski JA, et al. Estimates of Drug Overdose Deaths involving Fentanyl, Methamphetamine, Cocaine, Heroin, and Oxycodone: United States, 2021. 2023.

3 Pathan H, Williams J. Basic opioid pharmacology: an update. British journal of pain. 2012;6:11–16.

4 Handal KA, Schauben JL, Salamone FR. Naloxone. Annals of emergency medicine. 1983;12:438–445.

5 Boyer EW. Management of opioid analgesic overdose. New England Journal of Medicine. 2012;367:146–155.

6 Blanco C, Volkow ND. Management of opioid use disorder in the USA: present status and future directions. Lancet. 2019;393:1760–1772.

7 Johansson J, Hirvonen J, Lovro Z, et al. Intranasal naloxone rapidly occupies brain mu-opioid receptors in human subjects. Neuropsychopharmacology. 2019;44:1667–1673.

8 Volkow ND, Blanco C. Fentanyl and Other Opioid Use Disorders: Treatment and Research Needs. Am J Psychiatry. 2023;180:410–417.

9 Peng J, Sarkar S, Chang SL. Opioid receptor expression in human brain and peripheral tissues using absolute quantitative real-time RT-PCR. Drug and alcohol dependence. 2012;124:223–228.

10 Volkow ND, McLellan AT. Opioid abuse in chronic pain—misconceptions and mitigation strategies. New England Journal of Medicine. 2016;374:1253–1263.

11 Zubieta J-K, Dannals RF, Frost JJ. Gender and age influences on human brain mu-opioid receptor binding measured by PET. American Journal of Psychiatry. 1999;156:842–848.

12 Hirvonen J, Aalto S, Hagelberg N, et al. Measurement of central µ-opioid receptor binding in vivo with PET and [^11^C] carfentanil: a test–retest study in healthy subjects. European journal of nuclear medicine and molecular imaging. 2009;36:275–286.

13 Endres CJ, Bencherif B, Hilton J, Madar I, Frost JJ. Quantification of brain μ-opioid receptors with [11C] carfentanil: reference-tissue methods. Nuclear medicine and biology. 2003;30:177–186.

14 Tuominen L, Salo J, Hirvonen J, et al. Temperament trait harm avoidance associates with μ-opioid receptor availability in frontal cortex: a PET study using [11C] carfentanil. Neuroimage. 2012;61:670–676.

15 Heinz A, Reimold M, Wrase J, et al. Correlation of stable elevations in striatal μ-opioid receptor availability in detoxified alcoholic patients with alcohol craving: a positron emission tomography study using carbon 11–labeled carfentanil. Archives of general psychiatry. 2005;62:57–64.

16 Saccone PA, Lindsey AM, Koeppe RA, et al. Intranasal opioid administration in rhesus monkeys: PET imaging and antinociception. Journal of Pharmacology and Experimental Therapeutics. 2016;359:366–373.

17 Scott PJ, Koeppe RA, Shao X, et al. The effects of intramuscular naloxone dose on Mu receptor displacement of Carfentanil in Rhesus monkeys. Molecules. 2020;25:1360.

18 Kang Y, O’Conor KA, Kelleher AC, et al. Naloxone’s dose-dependent displacement of [^11^C] carfentanil and duration of receptor occupancy in the rat brain. Scientific Reports. 2022;12:6429.

19 Jalal H, Burke DS. Carfentanil and the rise and fall of overdose deaths in the United States. Addiction. 2021;116:1593–1599.

20 Zawilska JB, Kuczynska K, Kosmal W, Markiewicz K, Adamowicz P. Carfentanil - from an animal anesthetic to a deadly illicit drug. Forensic Sci Int. 2021;320:110715.

21 Frost JJ, Douglass KH, Mayberg HS, et al. Multicompartmental analysis of [11C]-carfentanil binding to opiate receptors in humans measured by positron emission tomography. J Cereb Blood Flow Metab. 1989;9:398–409.

22 DosSantos MF, Martikainen IK, Nascimento TD, et al. Reduced basal ganglia mu-opioid receptor availability in trigeminal neuropathic pain: a pilot study. Mol Pain. 2012;8:74.

23 Kim DJ, Nascimento TD, Lim M, et al. Exploring HD-tDCS Effect on mu-opioid Receptor and Pain Sensitivity in Temporomandibular Disorder: A Pilot Randomized Clinical Trial Study. J Pain. 2024;25:1070–1081.

24 Lamusuo S, Hirvonen J, Lindholm P, et al. Neurotransmitters behind pain relief with transcranial magnetic stimulation - positron emission tomography evidence for release of endogenous opioids. Eur J Pain. 2017;21:1505–1515.

25 Nascimento TD, Yang N, Salman D, et al. micro-Opioid Activity in Chronic TMD Pain Is Associated with COMT Polymorphism. J Dent Res. 2019;98:1324–1331.

26 Smith YR, Stohler CS, Nichols TE, Bueller JA, Koeppe RA, Zubieta JK. Pronociceptive and antinociceptive effects of estradiol through endogenous opioid neurotransmission in women. J Neurosci. 2006;26:5777–5785.

27 Jern P, Chen J, Tuisku J, et al. Endogenous Opioid Release After Orgasm in Man: A Combined PET/Functional MRI Study. J Nucl Med. 2023;64:1310–1313.

28 Karlsson HK, Tuominen L, Tuulari JJ, et al. Obesity is associated with decreased mu-opioid but unaltered dopamine D2 receptor availability in the brain. J Neurosci. 2015;35:3959–3965.

29 Majuri J, Joutsa J, Johansson J, et al. Dopamine and Opioid Neurotransmission in Behavioral Addictions: A Comparative PET Study in Pathological Gambling and Binge Eating. Neuropsychopharmacology. 2017;42:1169–1177.

30 Saanijoki T, Tuominen L, Tuulari JJ, et al. Opioid Release after High-Intensity Interval Training in Healthy Human Subjects. Neuropsychopharmacology. 2018;43:246–254.

31 Colasanti A, Searle GE, Long CJ, et al. Endogenous opioid release in the human brain reward system induced by acute amphetamine administration. Biol Psychiatry. 2012;72:371–377.

32 Domino EF, Hirasawa-Fujita M, Ni L, Guthrie SK, Zubieta JK. Regional brain [(11)C]carfentanil binding following tobacco smoking. Prog Neuropsychopharmacol Biol Psychiatry. 2015;59:100–104.

33 Gorelick DA, Kim YK, Bencherif B, et al. Imaging brain mu-opioid receptors in abstinent cocaine users: time course and relation to cocaine craving. Biol Psychiatry. 2005;57:1573–1582.

34 Mick I, Myers J, Stokes PR, et al. Amphetamine induced endogenous opioid release in the human brain detected with [(1)(1)C]carfentanil PET: replication in an independent cohort. Int J Neuropsychopharmacol. 2014;17:2069–2074.

35 Turton S, Myers JF, Mick I, et al. Blunted endogenous opioid release following an oral dexamphetamine challenge in abstinent alcohol-dependent individuals. Mol Psychiatry. 2020;25:1749–1758.

36 Zubieta JK, Dannals RF, Frost JJ. Gender and age influences on human brain mu-opioid receptor binding measured by PET. Am J Psychiatry. 1999;156:842–848.

37 Love T, Shabalin AA, Kember RL, et al. Unique and joint associations of polygenic risk for major depression and opioid use disorder with endogenous opioid system function. Neuropsychopharmacology. 2022;47:1784–1790.

38 Madar I, Bencherif B, Lever J, et al. Imaging delta- and mu-opioid receptors by PET in lung carcinoma patients. J Nucl Med. 2007;48:207–213.

39 Villemagne PS, Dannals RF, Ravert HT, Frost JJ. PET imaging of human cardiac opioid receptors. Eur J Nucl Med Mol Imaging. 2002;29:1385–1388.

40 Cherry SR, Jones T, Karp JS, Qi J, Moses WW, Badawi RD. Total-Body PET: Maximizing Sensitivity to Create New Opportunities for Clinical Research and Patient Care. J Nucl Med. 2018;59:3–12.

41 Dai B, Daube-Witherspoon ME, McDonald S, et al. Performance evaluation of the PennPET explorer with expanded axial coverage. Phys Med Biol. 2023;68.

42 Karp JS, Viswanath V, Geagan MJ, et al. PennPET Explorer: Design and Preliminary Performance of a Whole-Body Imager. J Nucl Med. 2020;61:136–143.

43 Pantel AR, Viswanath V, Daube-Witherspoon ME, et al. PennPET Explorer: Human Imaging on a Whole-Body Imager. J Nucl Med. 2020;61:144–151.

44 Viswanath V, Pantel AR, Daube-Witherspoon ME, et al. Quantifying bias and precision of kinetic parameter estimation on the PennPET Explorer, a long axial field-of-view scanner. IEEE Trans Radiat Plasma Med Sci. 2020;4:735–749.

45 Hsieh CJ, Hou C, Lee H, et al. Total-body imaging of mu-opioid receptors with [(11)C]carfentanil in non-human primates. Eur J Nucl Med Mol Imaging. 2024.

46 Sheehan DV, Lecrubier Y, Sheehan KH, et al. The Mini-International Neuropsychiatric Interview (M.I.N.I.): the development and validation of a structured diagnostic psychiatric interview for DSM-IV and ICD-10. J Clin Psychiatry. 1998;59 Suppl 20:22-33;quiz 34-57.

47 American Psychiatric Association., American Psychiatric Association. DSM-5 Task Force. Diagnostic and statistical manual of mental disorders : DSM-5. Fifth edition. ed.

48 Collins DL, Zijdenbos AP, Kollokian V, et al. Design and construction of a realistic digital brain phantom. IEEE Trans Med Imaging. 1998;17:463–468.

49 Tzourio-Mazoyer N, Landeau B, Papathanassiou D, et al. Automated anatomical labeling of activations in SPM using a macroscopic anatomical parcellation of the MNI MRI single-subject brain. Neuroimage. 2002;15:273–289.

50 Logan J, Fowler JS, Volkow ND, Wang GJ, Ding YS, Alexoff DL. Distribution volume ratios without blood sampling from graphical analysis of PET data. J Cereb Blood Flow Metab. 1996;16:834–840.

51 Zubieta JK, Gorelick DA, Stauffer R, Ravert HT, Dannals RF, Frost JJ. Increased mu opioid receptor binding detected by PET in cocaine-dependent men is associated with cocaine craving. Nat Med. 1996;2:1225–1229.

52 Czlonkowski A, Costa T, Przewlocki R, Pasi A, Herz A. Opiate receptor binding sites in human spinal cord. Brain Res. 1983;267:392–396.

53 Li EJ, Lopez JE, Spencer BA, et al. Total-Body Perfusion Imaging with [(11)C]-Butanol. J Nucl Med. 2023;64:1831–1838.

54 Cunningham VJ, Hume SP, Price GR, Ahier RG, Cremer JE, Jones AK. Compartmental analysis of diprenorphine binding to opiate receptors in the rat in vivo and its comparison with equilibrium data in vitro. J Cereb Blood Flow Metab. 1991;11:1–9.

55 Innis RB, Cunningham VJ, Delforge J, et al. Consensus nomenclature for in vivo imaging of reversibly binding radioligands. J Cereb Blood Flow Metab. 2007;27:1533–1539.

56 Zanderigo F, Ogden RT, Parsey RV. Reference region approaches in PET: a comparative study on multiple radioligands. J Cereb Blood Flow Metab. 2013;33:888–897.

57 Gunn RN, Lammertsma AA, Hume SP, Cunningham VJ. Parametric imaging of ligand-receptor binding in PET using a simplified reference region model. Neuroimage. 1997;6:279–287.

58 van der Weijden CWJ, Mossel P, Bartels AL, et al. Non-invasive kinetic modelling approaches for quantitative analysis of brain PET studies. Eur J Nucl Med Mol Imaging. 2023;50:1636–1650.

