## Supplemental Figures for "[^11^C]Carfentanil PET Whole-Body Imaging of Mu-Opioid Receptors: A First In-Human Study"

**Figure S1**

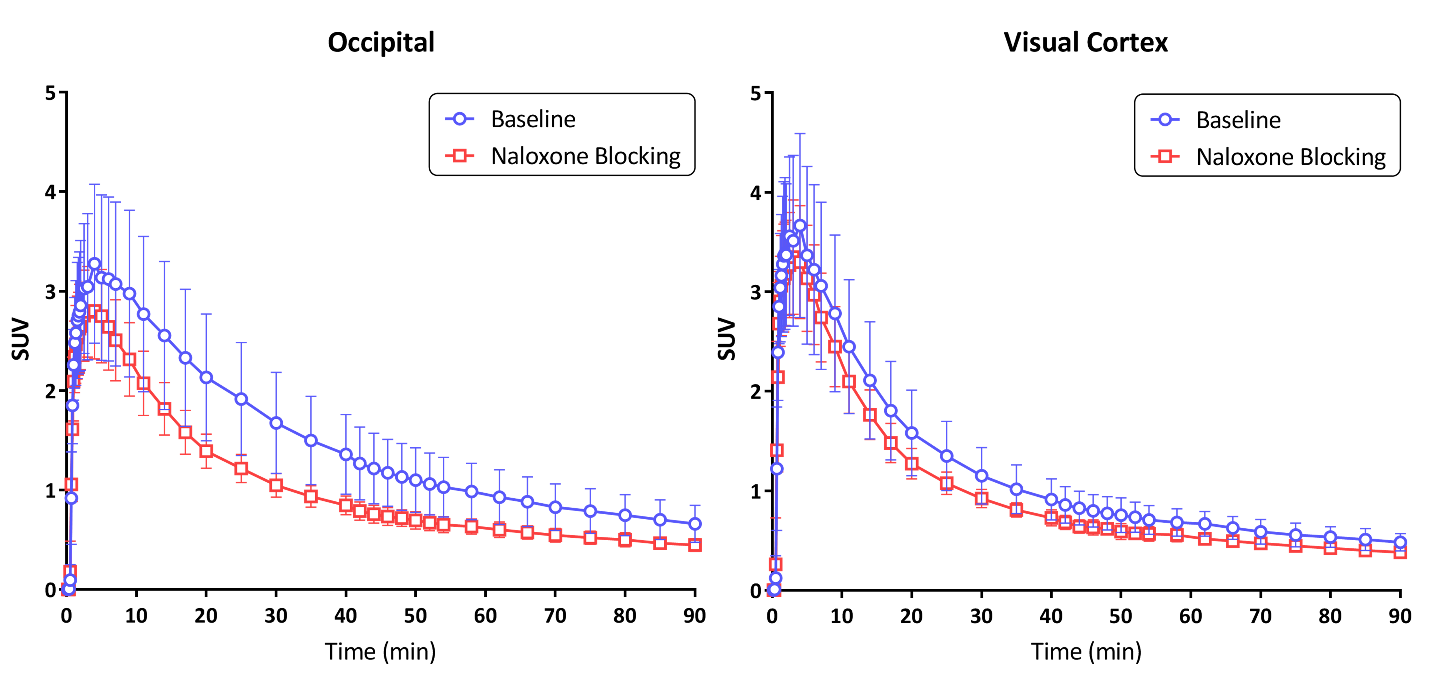

**Supplemental Figure S1. Brain reference region time activity curves.** Figure S1 shows differences in the time-activity curves (TACs) between baseline and naloxone for the occipital reference region, as defined by the AAL atlas (left panel). Because the ideal reference region is devoid of receptors, there should be no difference between the curves. When visual cortex (calcarine) is used as the reference region, no difference is observed (right panel).

**Figure S2**

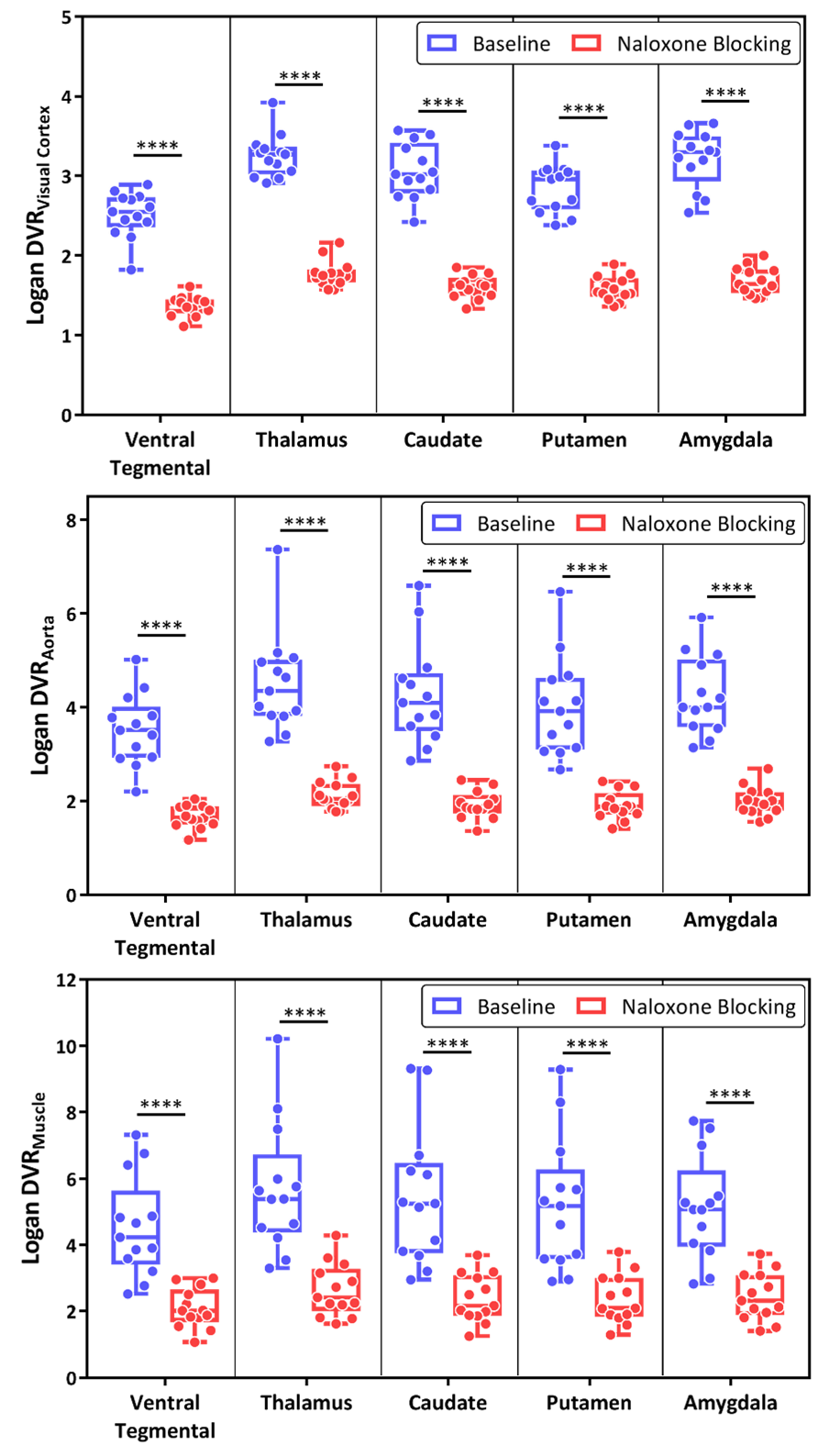

**Supplemental Figure S2. Using peripheral reference regions reiterates naloxone MOR antagonism.** Figure S2 shows the effect of different reference regions in determining MOR availability in MOR-rich brain regions: ventral tegmentum, thalamus, caudate, putamen, and amygdala. Reference regions include visual cortex (top panel), descending aorta (middle panel), and upper extremity muscle (bottom panel). Note that all three reference regions available using the LAFOV PET instrument demonstrate that naloxone reduces MOR availability.

**Figure S3**

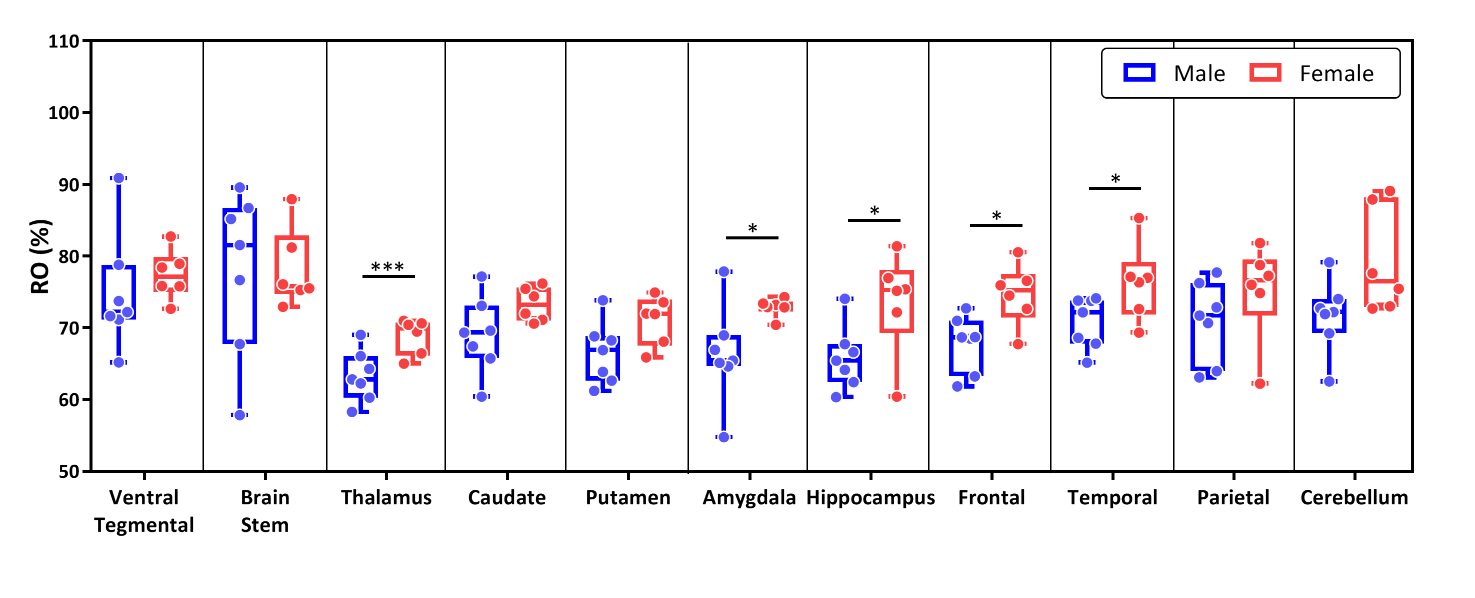

**Supplemental Figure S3.** **Naloxone MOR %RO throughout the brain.**  The top panel of supplemental figure S3 shows sex differences in the naloxone receptor occupancy of MORs. Unpaired t-test assuming unequal variances (* p<0.05, *** p<0.001). Note the differences in the frontal and temporal cortices and the hippocampus, as well as the MOR-rich regions of thalamus and amygdala.

**Figure S4**

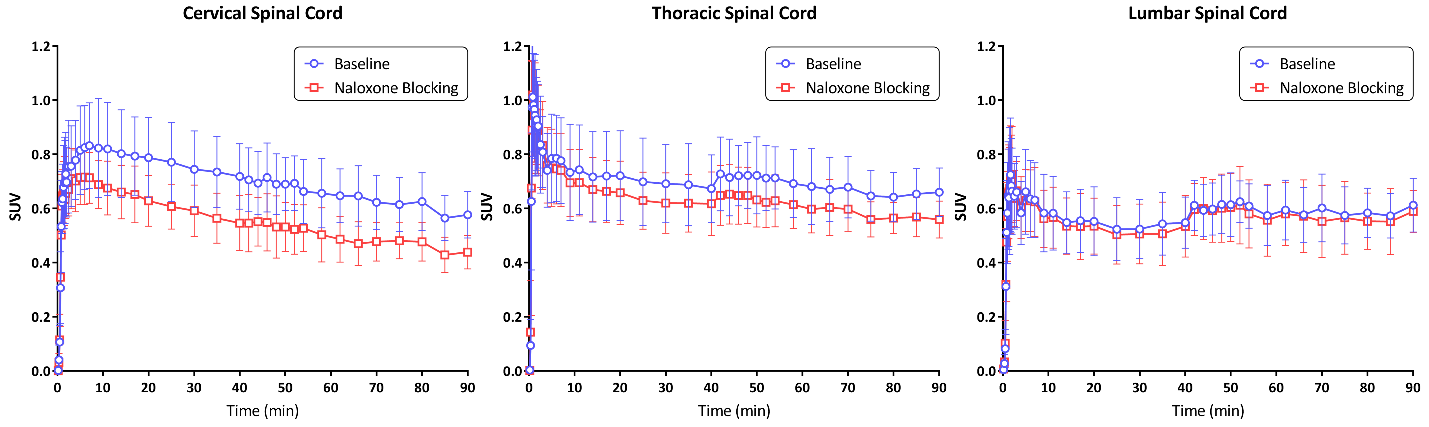

**Supplemental Figure S4. Spinal Cord Time Activity Curves (TACs)**. SUV TACs for the cervical (left panel), thoracic (middle panel), and lumbar (right panel) portions of the spinal cord are shown in figure S4. Note the separation between baseline and naloxone blocking conditions for the cervical and thoracic portions of the spinal cord.

**Supplemental Table T1: Brain region DVRs (visual cortex reference region) at baseline and post naloxone block; naloxone MOR %RO (mean data value +/-standard deviation).**

|  | Male (n=7) | | | Female (n=6) | | |
| --- | --- | --- | --- | --- | --- | --- |
|  | Baseline | Naloxone Blocking | %RO | Baseline | Naloxone Blocking | %RO |
| Ventral Tegmental | 2.5 ± 0.4 | 1.4 ± 0.2 | 74.8 ± 8.1 | 2.5 ± 0.2 | 1.3 ± 0.1 | 77.4 ± 3.5 |
| Brain Stem | 2.0 ± 0.1 | 1.2 ± 0.2 | 77.9 ± 11.5 | 1.9 ± 0.2 | 1.2 ± 0.1 | 78.2 ± 5.5 |
| Thalamus | 3.4 ± 0.3 | 1.9 ± 0.2 | 63.3 ± 3.6 | 3.1 ± 0.2 | 1.7 ± 0.1 | 68.8 ± 2.5 |
| Caudate | 3.1 ± 0.3 | 1.7 ± 0.2 | 69.0 ± 5.3 | 3.1 ± 0.4 | 1.5 ± 0.1 | 73.3 ± 2.4 |
| Putamen | 2.9 ± 0.3 | 1.7 ± 0.2 | 66.5 ± 4.3 | 2.8 ± 0.3 | 1.5 ± 0.1 | 71.1 ± 3.4 |
| Amygdala | 3.3 ± 0.3 | 1.8 ± 0.2 | 66.2 ± 6.8 | 3.2 ± 0.4 | 1.6 ± 0.1 | 72.8 ± 1.3 |
| Hippocampus | 1.8 ± 0.1 | 1.3 ± 0.0 | 65.8 ± 4.4 | 1.8 ± 0.1 | 1.2 ± 0.0 | 73.6 ± 7.1 |
| Frontal | 2.0 ± 0.2 | 1.3 ± 0.1 | 67.8 ± 3.9 | 1.9 ± 0.2 | 1.2 ± 0.1 | 74.7 ± 4.3 |
| Temporal | 2.0 ± 0.3 | 1.3 ± 0.1 | 70.8 ± 3.6 | 1.9 ± 0.2 | 1.2 ± 0.1 | 76.3 ± 5.4 |
| Parietal | 1.7 ± 0.1 | 1.2 ± 0.0 | 70.9 ± 5.6 | 1.6 ± 0.1 | 1.2 ± 0.0 | 75.2 ± 6.8 |
| Cerebellum | 2.0 ± 0.2 | 1.3 ± 0.1 | 71.7 ± 5.0 | 1.7 ± 0.2 | 1.2 ± 0.1 | 79.3 ± 7.4 |

**Supplemental Table T2: Whole-body VOIs determined using SUVR_Aorta_ values summed 50-70 minutes following [^11^C]CFN administration. P-value determined using unpaired t-test assuming unequal variances.**

|  | All (n=13) | |  | Male (n=7) | |  | Female (n=6) | |  |
| --- | --- | --- | --- | --- | --- | --- | --- | --- | --- |
|  | Baseline | Naloxone  Blocking | p-value | Baseline | Naloxone  Blocking | p-value | Baseline | Naloxone  Blocking | p-value |
| Heart Wall | 2.71 ± 0.91 | 2.65 ± 0.66 | 0.6140 | 2.98 ± 1.09 | 2.83 ± 0.74 | 0.4614 | 2.39 ± 0.58 | 2.44 ± 0.55 | 0.7149 |
| Kidney Cortex | 3.57 ± 0.40 | 3.57 ± 0.44 | 0.9826 | 3.80 ± 0.33 | 3.84 ± 0.39 | 0.8131 | 3.29 ± 0.29 | 3.25 ± 0.24 | 0.7321 |
| Spleen | 3.10 ± 0.57 | 3.01 ± 0.42 | 0.4149 | 3.11 ± 0.25 | 2.97 ± 0.31 | 0.3897 | 3.07 ± 0.84 | 3.06 ± 0.54 | 0.9026 |
| Small Bowel | 7.81 ± 1.61 | 7.42 ± 1.18 | 0.9378 | 8.56 ± 1.36 | 7.93 ± 1.14 | 0.5708 | 6.94 ± 1.52 | 6.83 ± 0.99 | 0.5664 |
| C-Spinal Bone  Marrow | 5.25 ± 0.96 | 4.65 ± 0.94 | ***0.0096*** | 5.46 ± 0.81 | 4.53 ± 0.86 | ***0.0111*** | 5.02 ± 1.14 | 4.80 ± 1.08 | 0.3808 |
| T-Spinal Bone  Marrow | 6.18 ± 1.07 | 5.85 ± 0.89 | 0.2118 | 6.16 ± 0.94 | 5.74 ± 0.63 | 0.3300 | 6.19 ± 1.30 | 5.98 ± 1.18 | 0.5095 |
| L-Spinal Bone  Marrow | 6.26 ± 1.06 | 5.93 ± 1.08 | 0.1981 | 6.34 ± 1.01 | 5.85 ± 1.03 | 0.2589 | 6.18 ± 1.20 | 6.03 ± 1.22 | 0.6226 |
| C-Spinal Cord | 1.17 ± 0.21 | 0.91 ± 0.13 | ***0.0027*** | 1.21 ± 0.24 | 0.89 ± 0.17 | ***0.0227*** | 1.12 ± 0.16 | 0.94 ± 0.06 | 0.0727 |
| T-Spinal Cord | 1.24 ± 0.19 | 1.12 ± 0.14 | ***0.0111*** | 1.20 ± 0.21 | 1.07 ± 0.15 | 0.0547 | 1.30 ± 0.16 | 1.17 ± 0.10 | 0.1437 |
| L-Spinal Cord | 1.09 ± 0.15 | 1.05 ± 0.19 | 0.3791 | 1.10 ± 0.16 | 1.03 ± 0.23 | 0.5074 | 1.09 ± 0.15 | 1.06 ± 0.16 | 0.5762 |
| Muscle | 0.98 ± 0.18 | 1.04 ± 0.20 | 0.2679 | 0.96 ± 0.14 | 0.96 ± 0.16 | 0.9026 | 1.01 ± 0.23 | 1.13 ± 0.22 | 0.1917 |
| Liver | 14.67 ± 3.15 | 14.65 ± 2.40 | 0.9762 | 15.36 ± 3.66 | 15.02 ± 2.67 | 0.7723 | 13.87 ± 2.53 | 14.23 ± 2.20 | 0.2725 |
| Stomach | 11.35 ± 7.53 | 11.64 ± 12.04 | 0.4431 | 10.64 ± 4.96 | 13.82 ± 16.35 | 0.4370 | 12.18 ± 10.24 | 9.10 ± 3.55 | 0.8734 |

**Supplemental Table T3: Area Under Curve (AUC) summation for reference regions (occipital cortex, visual cortex, descending aorta, and upper extremity muscle) at baseline and after naloxone block. P-value determined using unpaired t-test assuming unequal variances.**

| Region | Baseline | Naloxone Blocking | p-value |
| --- | --- | --- | --- |
| Occipital Cortex | 132.3 ± 35.5 | 91.4 ± 11.1 | 0.0002 |
| Visual Cortex | 105.5 ± 23.5 | 87.7 ± 9.9 | 0.0020 |
| Aorta | 66.8 ± 7.3 | 64.7 ± 4.9 | 0.4059 |
| Muscle | 48.0 ± 15.1 | 47.7 ± 11.7 | 0.9104 |
| Cervical Spinal Cord | 62.1 ± 9.9 | 48.2 ± 6.0 | <0.0001 |
| Thoracic Spinal Cord | 62.4 ± 11.7 | 55.2 ± 7.9 | 0.0039 |
| Lumbar Spinal Cord | 51.3 ± 8.5 | 48.6 ± 9.2 | 0.1921 |
